# A multinational pharmacoepidemiology atlas of prescription medication associations with all-cause mortality across six population cohorts

**DOI:** 10.64898/2026.11.12.26367024

**Authors:** Kevin Perez, Jonathan K. L. Mak, Kathryn C. B. Tan, Andrea Ganna, Alessandro Ceschi, Juulia Jylhävä, Ching-Lung Cheung, Andrea Alimonti

## Abstract

Observational studies sometimes report that individual medications are associated with lower mortality, but such signals are rarely compared across drugs and health systems. We screened prescription medications for associations with all-cause mortality using time-dependent Cox models in six cohorts from the UK, US, Finland, Hong Kong and Japan (8.93 million individuals aged 40 years or older; 1.49 billion prescriptions). Most medications were associated with higher mortality, reflecting confounding by indication. A few medications showed inverse associations with mortality, notably the varicella-zoster virus (VZV) vaccine, the antimalarials atovaquone-proguanil (Malarone) and mefloquine, and the hormone replacement therapy estradiol. These associations were only modestly attenuated by adjustment for comorbidity, lifestyle and socio-economic factors, and persisted within strata defined by pre-existing disease. While adjustment for self-reported health abolished the Malarone association, probably reflecting a healthy traveler profile, the VZV association was maintained. In FinnGen, deaths from external causes, which vaccination cannot prevent, were lower among VZV vaccine users, but much less so than all-cause mortality. In a target trial emulation in All of Us, the VZV association was maintained against matched unvaccinated individuals (HR 0.61, 95% CI 0.53-0.70) and pneumococcal vaccine recipients (HR 0.64, 0.55-0.74), but was attenuated against recipients of tetanus-diphtheria vaccines, a group that also has low mortality (HR 0.85, 0.70-1.03). Healthy-user selection contributes to these signals but does not appear to fully explain the VZV vaccine association, which warrants confirmation in randomized trials. This multinational atlas provides a resource for prioritizing drug-repurposing candidates in ageing research.

## INTRODUCTION

Population ageing has intensified interest in whether existing medications influence survival beyond their primary indication^1,2^. Observational studies have reported such associations for individual drugs^3^, but each typically examines a single agent in a single health system, with study-specific designs and covariates. This makes it difficult to judge whether a given signal is reproducible, or how it compares with the many medications never examined. Several such signals have later failed to replicate in randomized trials, hormone replacement therapy being the best-known example^4,5^, largely because of confounding by indication, immortal time bias and healthy-user bias^6–8^. Real-world data (RWD) from electronic health records and population biobanks now make it possible to examine many medications simultaneously^9^. Such a screen cannot establish causal effects, but it can serve three purposes: mapping drug–mortality associations under a single pipeline; testing whether specific signals replicate across health systems with different prescribing practices; and characterizing how sensitive each signal is to adjustment for comorbidity, lifestyle and self-reported health, that is, how much of it is plausibly explained by who receives the drug.

To address this knowledge gap, we developed a multinational pharmacoepidemiology framework to systematically evaluate the association between prescription drug utilization and all-cause mortality, leveraging six independent cohorts spanning three continents. Our investigation integrates harmonized longitudinal data from the All of Us^10^ (AOU) Research Program and the National Health and Nutrition Examination Survey^11^ (NHANES) in the United States; the UK Biobank^12^ (UKBB) and FinnGen^13^ in Europe; alongside the Hong Kong territory-wide EHR database (Clinical Data Analysis and Reporting System^14^; CDARS) and the Japan Medical Data Center^15^ (JMDC) claims database in Asia. This trans-continental integration captures a cross-section of global prescribing practices, sociodemographic profiles, and healthcare infrastructures.

To examine survival outcomes while mitigating temporal biases, we applied time-dependent covariate^16^ (TDC) Cox proportional hazards models across the prescribed drug catalog. We utilized a stepwise adjustment pipeline: beginning with baseline demographic adjustments (age, sex and enrollment year), followed by clinical risk adjustments utilizing the Charlson Comorbidity Index^17^ (CCI), and concluding with adjustments for lifestyle and socioeconomic factors, including smoking status, body mass index (BMI) and socio-economic status (SES). Lastly, to determine whether increased survival is maintained in different clinical contexts, we conducted stratified analyses across sex and major clinical subgroups (e.g., isolating effects within oncology or diabetology cohorts versus relatively healthy baselines). Finally, to probe healthy-user bias directly, we examined a negative-control exposure that shares the selection profile of a candidate drug but has no plausible effect on mortality, and cause-specific mortality outcomes that the drugs cannot plausibly affect. For the VZV vaccine, we additionally emulated a target trial^18^, comparing vaccinees with unvaccinated individuals and with recipients of other adult vaccines.

We present the resulting atlas as an openly available, hypothesis-generating resource to help prioritize candidates for drug repurposing and further study. Its associations are not estimates of causal drug effects: inverse associations ultimately require drug-specific randomized trials, before they can inform practice, and importantly, associations with higher mortality should not be read as evidence of harm.

## RESULTS

### Cohort Description

We harmonized longitudinal data from six population cohorts, together encompassing 8.93 million individuals and 1.49 billion prescriptions (**Table 1**). The cohorts differed widely in design and clinical profile, spanning population biobanks with deep phenotyping (UKBB, FinnGen, AOU), a nationally representative survey (NHANES), a territory-wide hospital database of older adults (CDARS) and an insurance claims database of working-age adults and their dependants (JMDC). Accordingly, mean age at enrollment ranged from 52 years in JMDC to 68 years in CDARS, median follow-up from 3.4 years in AOU to 14.7 years in UKBB, and the proportion of participants who died during follow-up from 0.4% in JMDC to 29.5% in CDARS. This heterogeneity in populations, healthcare systems and data capture provides a stringent test of whether drug–mortality associations replicate across settings.

**Table 1.**
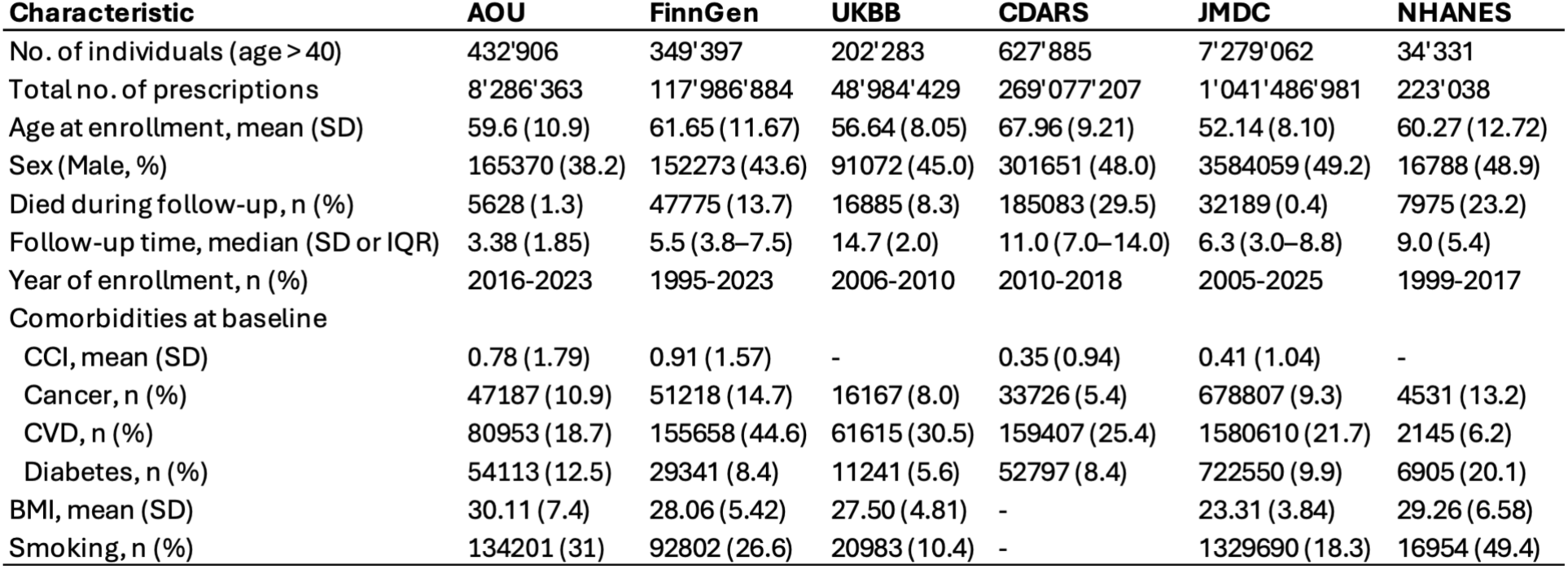
Baseline demographics and clinical characteristics of the six population cohorts. Demographic profiles, follow-up, mortality and prescription volumes for the 8.93 million individuals included (UKBB, NHANES, AOU, FinnGen, CDARS and JMDC). Baseline comorbidities include the Charlson Comorbidity Index (CCI), cancer, circulatory disease (CVD; ICD-10 I00–I99), diabetes, body mass index (BMI) and smoking status. Continuous variables are presented as means (SD) or medians (IQR), and categorical variables as counts (%).

### Drugs Associated with Lower All-Cause Mortality

To capture the dynamic nature of drug exposure and minimize immortal time bias, we employed a time-dependent covariate (TDC) survival analysis utilizing Cox proportional hazards models (**Figure 1, S1**). This framework allowed us to evaluate the hazard ratios (HR) for all-cause mortality associated with specific prescription medications with progressive adjustment for baseline comorbidities, in subjects aged 40yo or older. We first evaluated and meta-analyzed these associations in a first model (Model 1), adjusted for sex, age and year of enrollment. Pooled estimates combined AOU, UKBB, FinnGen and CDARS; NHANES, which only captured medication use at baseline, and JMDC, which comprised over 80% of all participants and would dominate pooled estimates, are shown at the cohort level but were not pooled (Methods).

**Figure 1.**
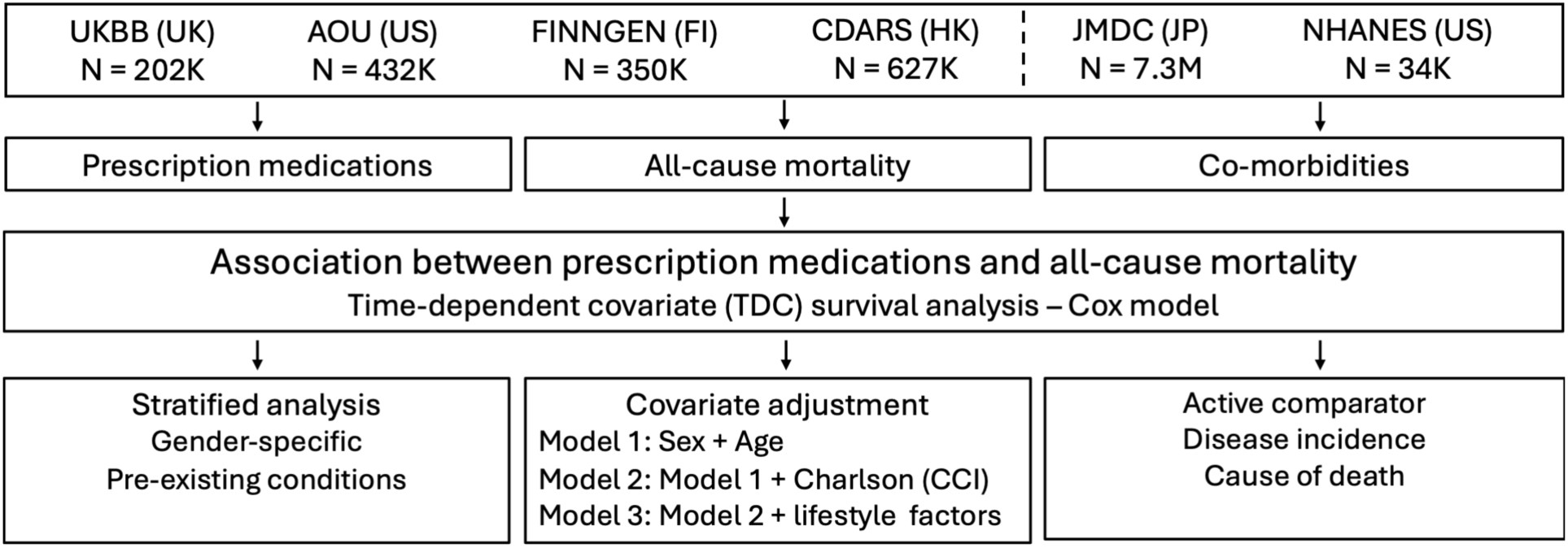
Study design and analytical framework. Overview of the multinational pharmacoepidemiology study evaluating the association between prescription medications and all-cause mortality. The study integrated and harmonized longitudinal data from six population cohorts: UK Biobank (UKBB), All of Us (AOU), National Health and Nutrition Examination Survey (NHANES), Japan Medical Data Center (JMDC), Clinical Data Analysis and Reporting System (CDARS) and FinnGen. A time-dependent covariate (TDC) survival analysis using Cox proportional hazards models was employed to mitigate immortal time bias. The analytical pipeline included progressively stringent covariate adjustments (Models 1–3), stratified analyses by sex and pre-existing clinical conditions, and sensitivity analyses (active comparators, negative-control exposure and outcomes).

In this model, most medications were associated with higher mortality (HR > 1; **Figure 2A**). This is the expected pattern when users are compared with all non-users: medications are prescribed to people with the conditions they treat, and those conditions, rather than the drugs, carry the excess risk (confounding by indication). These HRs should therefore not be interpreted as evidence of harm. Against this background, a small number of medications were associated with lower mortality across cohorts. These include notably the Shingles (VZV) vaccine, the DTP vaccine and the antimalarials atovaquone/proguanil (Malarone) and mefloquine, which remained significant after Bonferroni correction (**Figure 2B**). Typhoid vaccine (HR 0.68, 95% CI 0.43–1.09), tibolone (HR 0.71, 0.44–1.15) and estradiol (HR 0.77, 0.63–0.94) also showed inverse associations of similar magnitude. To further characterize these associations, we conducted sex-specific analyses adjusted for baseline age and enrollment year (“Model 1”), which showed that the lower mortality association for most medications was maintained across both male and female populations (**Figure 2C**). Lower mortality, sex-agnostic signals were observed for prophylactic immunizations, including the VZV (varicella-zoster) vaccine and the DTP vaccine, as well as the antimalarial combination of atovaquone/proguanil alongside mefloquine. Hormone therapies (estradiol, tibolone) were, as expected, prescribed almost exclusively to women.

**Figure 2.**
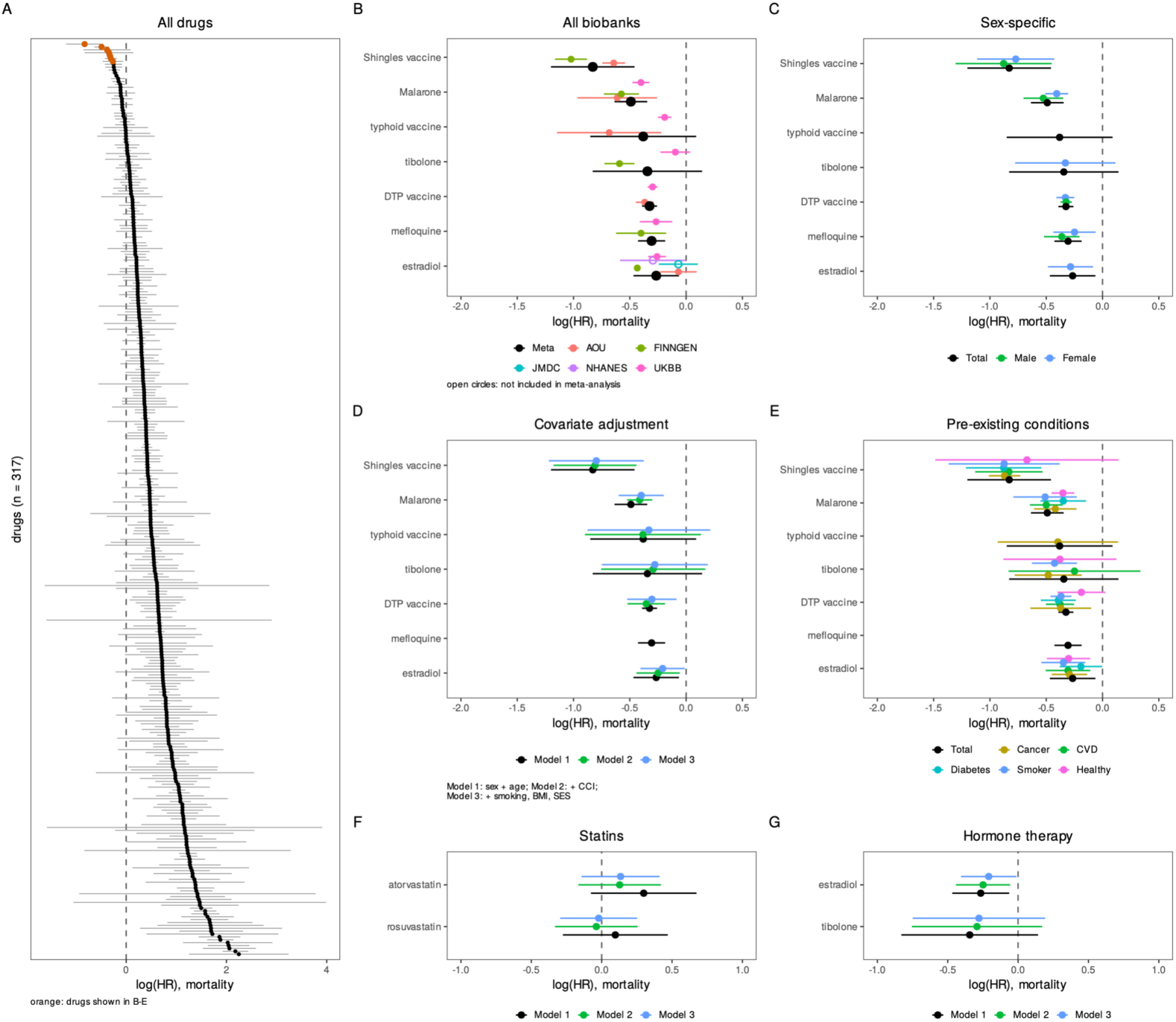
Drugs associated with lower all-cause mortality across adjustment models and clinical strata. (A) Pooled mortality hazard ratios (HR) and 95% confidence intervals (CI) for all meta-analyzed drugs (Model 1; n = 317); drugs shown in B–E are highlighted in orange. HRs above 1 predominantly reflect confounding by indication, as users are compared with a reference group that largely lacks the treated condition, and should not be interpreted as evidence of harm. (B) HR by cohort (colors) and pooled estimate (black) for the drugs with the lowest pooled HR, down to estradiol (drug combinations excluded, except atovaquone/proguanil [Malarone]). JMDC and NHANES (open circles) are shown but were not included in the meta-analysis. (C) Total, male and female analyses (Model 1). (D) Adjustment models: Model 1 (sex, age and year of enrollment), Model 2 (+ CCI) and Model 3 (+ smoking, BMI and socio-economic status). (E) Analyses stratified by pre-existing cancer, circulatory disease (CVD) or diabetes, smoking, and in a healthy sub-cohort free of these conditions (Model 1). (F, G) Statins (atorvastatin, rosuvastatin) and hormone therapy (estradiol, tibolone) across Models 1–3. Pooled estimates are random-effects (REML) meta-analyses of AOU, UKBB, FinnGen and CDARS, computed when a drug was analyzed in at least two of these cohorts; missing points indicate fewer than two cohorts. Data are presented as log(HR) with 95% CI.

To reduce confounding factors, we applied further adjustments to our base model (**Figure 2D**). We then looked at mortality associations adjusted for baseline CCI (“Model 2”), as well as lifestyle and socioeconomic factors (“Model 3”), including smoking status, BMI, and SES, measured using the Townsend deprivation index (UK Biobank only). With Model 2 and Model 3, the inverse associations were only modestly attenuated (Shingles vaccine HR 0.44 in Model 1 and 0.45 in Model 3; DTP vaccine 0.72 and 0.74; Malarone 0.61 and 0.67; estradiol 0.77 and 0.81). In contrast, atorvastatin and rosuvastatin, statins that reduce all-cause mortality in randomized trials^19^, were not associated with lower mortality in any model, and adjustment for comorbidity moved their HRs from above 1 towards the null (atorvastatin 1.35 to 1.14; rosuvastatin 1.10 to 0.98; **Figure 2F**), consistent with confounding by indication rather than an absence of benefit. Estimates for estradiol and tibolone were essentially unchanged across models (**Figure 2G**). To determine if these lower mortality associations persisted within inherently high-risk or biologically vulnerable patient subpopulations, we conducted stratified analyses based on specific pre-existing clinical conditions, including cancer, circulatory disease (CVD), and diabetes, and behavioral stratification based on smoking status (**Figure 2E**). Across all these clinical subgroups, the pooled hazard ratios for the medications with the strongest inverse associations indicated lower mortality compared to non-users, with similar magnitudes in subgroups with very different baseline risks (e.g., Malarone HR 0.60–0.71 across strata). We also evaluated these medications within a relatively "healthy" patient sub-cohort, devoid of such comorbidities. Within this relatively healthier stratum, the inverse associations were generally maintained; for the Shingles vaccine, the pooled estimate became imprecise (HR 0.51, 95% CI 0.23–1.15).

Lastly, to reduce confounding by indication and healthy-user bias, we compared the medications with the strongest inverse associations against active comparators sharing their indication or preventive context (selection criteria in Methods; **Figure S2**). Compared with other adult vaccines (influenza, pneumococcal), the VZV and DTP vaccines remained associated with lower mortality. Estradiol was associated with lower mortality relative to estriol, but not relative to tibolone or conjugated estrogens.

### Probing Healthy-User Bias

We next examined whether the strongest inverse associations could reflect the characteristics of drug users rather than drug effects (**Figure 3**). First, in AOU, we additionally adjusted Model 1 for self-rated health, a strong predictor of mortality^20^. Malarone users reported markedly better health than users of the DTP and Shingles vaccines and than the overall cohort (**Figure 3A**), and adjustment attenuated the Malarone association towards the null, whereas the Shingles and DTP vaccine associations were largely maintained (**Figure 3B**).

**Figure 3.**
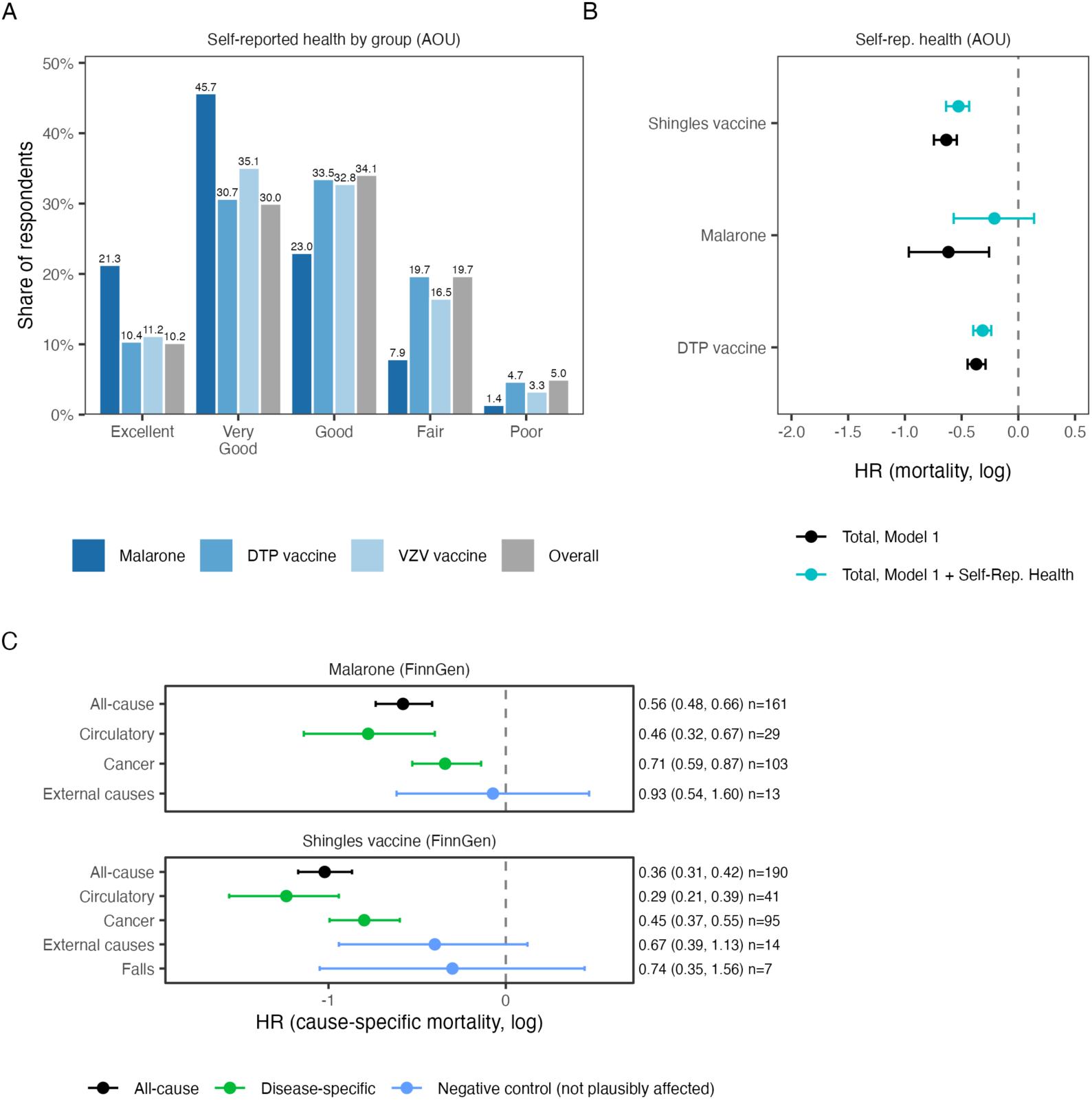
Probing healthy-user bias. (A) Distribution of self-rated health among users of atovaquone/proguanil (Malarone), the DTP vaccine and the Shingles (VZV) vaccine, and in the overall AOU cohort. (B) Mortality HR (95% CI) in AOU before (Model 1) and after additional adjustment for self-rated health. (C) Cause-specific mortality in FinnGen among users of Malarone and the Shingles vaccine: HR (95% CI) for death from all causes, circulatory disease (ICD-10 I00–I99), cancer (C00–D48), external causes (V01–Y98) and falls (W00–W19); external causes and falls serve as negative-control outcomes not plausibly affected by the drugs. Time-dependent Cox models (Model 1), with deaths from other causes censored. n, deaths among exposed; causes with fewer than five deaths among exposed are not shown.

Second, we used typhoid vaccine as a negative-control exposure for Malarone. Typhoid vaccine is given to the same population of international travelers but has no plausible effect on all-cause mortality in high-income settings. It was nevertheless associated with lower mortality (HR 0.68, 95% CI 0.43–1.09; AOU 0.51, UKBB 0.83), of similar magnitude to Malarone (HR 0.61, 0.53–0.71), and the association persisted after full adjustment (Model 3 HR 0.72; **Figure 2B, D**). This suggests that the Malarone association largely reflects the characteristics of travelers rather than a pharmacological effect.

Third, in FinnGen, where the underlying cause of death is available from the national registry, we estimated cause-specific mortality, treating deaths from other causes as censored (**Figure 3C**). Deaths from external causes (ICD-10 V01–Y98), which neither drug can plausibly prevent, served as negative-control outcomes. Among Shingles vaccine users, mortality from external causes (HR 0.67, 95% CI 0.39–1.13) and from falls (HR 0.74, 0.35– 1.56) was also lower, consistent with some degree of healthy-user selection. These associations were, however, considerably weaker than that for all-cause mortality (HR 0.36, 0.31–0.42), indicating that the selection captured by these outcomes does not by itself account for the full association. Circulatory (HR 0.29, 0.21–0.39) and cancer mortality (HR 0.45, 0.37–0.55) were also lower among users. Of note, the external-cause estimates rest on few events (14 and 7 deaths among users), so residual confounding by frailty cannot be excluded. For Malarone, mortality from external causes was not reduced (HR 0.93, 0.54– 1.60), whereas circulatory (HR 0.46, 0.32–0.67) and cancer mortality (HR 0.71, 0.59–0.87) were.

Finally, we emulated a target trial of VZV vaccination in AOU (Methods; **Figure S3, S4**). Compared with matched unvaccinated individuals, VZV vaccination remained associated with lower mortality (HR 0.61, 95% CI 0.53–0.70), as it did against pneumococcal vaccine recipients (HR 0.64, 0.55–0.74), a group often vaccinated because of chronic disease. Against Td/Tdap vaccine recipients, the most comparable group (self-rated health was balanced before weighting and all covariates after weighting), the association was attenuated (HR 0.85, 0.70–1.03). In all three comparisons, cumulative mortality curves separated soon after vaccination (**Figure S4**), with the strongest association in the first 90 days (e.g. HR 0.37, 0.21–0.65 against unvaccinated individuals), when a vaccine is unlikely to prevent death. Such early separation is a recognized sign of healthy-vaccinee selection^21^ and suggests that part of the VZV association may reflect who is vaccinated rather than an effect of the vaccine.

### Incident Age-Related Disease and Cause of Death

Lower all-cause mortality does not by itself indicate a delay in the onset of chronic disease. We therefore extended the TDC framework to three incident non-fatal outcomes — cancer, diabetes, and heart disease — modelled in parallel with all-cause mortality for the medications most associated with lower mortality: the VZV vaccine, the antimalarial combination atovaquone/proguanil, and estradiol (**Figure 4A**). Consistent with the primary analyses, users of all three agents showed lower cumulative mortality over five years of follow-up than non-users. The disease-specific outcomes, however, were heterogeneous. Incident diabetes was substantially lower among users of the VZV vaccine and Malarone, while the cumulative incidence of heart disease was essentially superimposable between users and non-users for all three medications. In contrast, incident cancer was consistently higher among users than non-users across all three agents. To determine whether this excess reflected genuine risk or differential ascertainment, we decomposed the composite cancer endpoint into its most frequent constituent diagnoses in AOU (**Figure 4B**). The excess was mainly composed of higher rates of basal cell carcinoma (BCC), and prostate cancer. Since these cancers have relatively lower mortality rates, this result is still compatible with the lower mortality rates observed. This pattern may also be consistent with a greater healthcare engagement, and higher screening uptake in these populations. Finally, in FinnGen, the distribution of causes of death among users shifted towards cancer and away from heart disease (**Figure 4C**). Because these are proportions of deaths, this shift reflects fewer circulatory deaths rather than more cancer deaths, consistent with the cause-specific analysis (**Figure 3C**). Taken together, users of the Shingles vaccine and Malarone had an incidence of heart disease similar to non-users in AOU, but markedly lower circulatory mortality in FinnGen.

**Figure 4.**
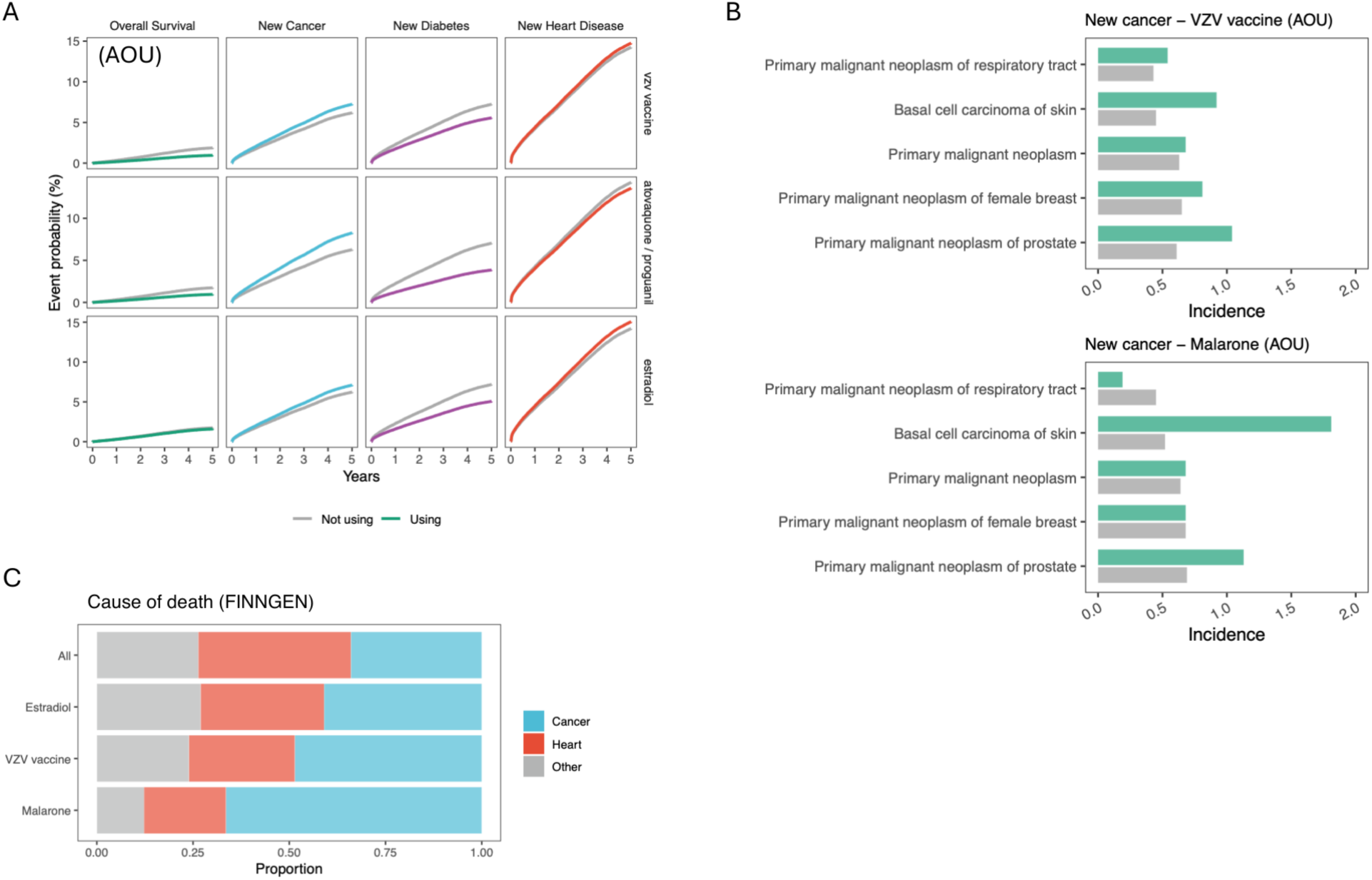
Associations of selected medications with incident age-related disease and cause of death. (A) Adjusted cumulative event probability (%) over five years for four outcomes (columns: all-cause mortality, incident cancer, incident diabetes, incident heart disease) with (colored) and without (grey) exposure to three medications (rows: VZV vaccine, atovaquone/proguanil, estradiol) in AOU. Curves are estimated from time-dependent Cox models adjusted for age and sex, for a woman of mean age. (B) Incidence of the five most frequently recorded cancer diagnoses among users (green) and non-users (grey) of atovaquone/proguanil (Malarone) and the VZV vaccine in AOU, per 100 subjects. (C) Distribution of underlying causes of death (cancer, heart disease, other) among all deceased participants and among deceased users of estradiol, the VZV vaccine and Malarone in FinnGen. Proportions describe the composition of deaths, not the risk of death; cause-specific mortality is shown in Figure 3C.

## Discussion

In this large-scale, multi-national analysis, we systematically evaluated the association between prescription medications and all-cause mortality across six diverse population-based cohorts. Our results identify several pharmacological classes associated with lower all-cause mortality, building upon and extending our previous findings that evaluated prescribed medications in the UK Biobank^22^. The current investigation deployed a time-dependent covariate (TDC) framework to minimize immortal time bias and capture the dynamic nature of drug exposure. Despite these methodological differences, our current cross-national analysis confirms the previously observed lower-mortality associations for medications such as estradiol, replicating these associations across multiple, independent healthcare systems and demographics. By combining six cohorts from three continents in a common analytical pipeline, and by probing healthy-user bias with complementary negative-control analyses, this work provides a framework for distinguishing robust drug– mortality signals from selection effects.

Two features of the design are important for interpreting the atlas. First, because each medication is compared with all non-users, many associations reflect confounding by indication. HRs above 1 largely index the severity of the treated condition, and the absence of an inverse association does not imply absence of benefit: statins, which reduce all-cause mortality in randomized trials^19^, appear null here. Second, the inverse associations that stand out against this background are concentrated in preventive and elective treatments (vaccines, travel prophylaxis and hormone therapy), the categories most susceptible to healthy-user bias^8^. The atlas should therefore be read as a ranked set of signals, together with their sensitivity to adjustment, rather than as estimates of drug effects.

A well-recognized challenge in observational pharmacoepidemiology is healthy-user bias^8^, wherein patients prescribed certain preventive therapies generally exhibit healthier baseline behaviors, better healthcare access, and higher socioeconomic status. For example, the antimalarial combination atovaquone/proguanil (Malarone), associated with lower mortality, may reflect a "healthy traveler effect," as individuals receiving prophylactic travel medications inherently possess the physical capacity, financial resources, and overall health required to undertake international travel. Other medications may also be subject to this bias, including vaccines and HRT. We probed this bias through stepwise adjustment for the CCI, lifestyle factors and self-reported health, and through stratification within highly morbid populations. These attenuated but did not abolish the associations. Still, residual healthy-user bias cannot be excluded. Some additional observations support this interpretation. Typhoid vaccine, which shares the traveler profile of Malarone but cannot plausibly reduce mortality, showed an inverse association of similar magnitude; Malarone users reported better self-rated health, and adjustment for it attenuated the Malarone association. For the Shingles vaccine, deaths from external causes and falls, which the vaccine cannot prevent, were also lower among users, but much less so than all-cause mortality. In a target trial emulation in AOU, the association was maintained against unvaccinated individuals and pneumococcal vaccine recipients but attenuated against recipients of Td/Tdap vaccines, and mortality curves separated soon after vaccination, when a protective effect is unlikely. The inverse association of the DTP vaccine itself points the same way. Yet when calibrated against DTP or Td/Tdap vaccines, which share this selection, a modest VZV association of roughly 15–25% lower mortality remained, compatible with a smaller true effect. Healthy-user selection therefore contributes to this association, but the residual association could reflect a true protective effect, which warrants evaluation in randomized trials.

Hormone replacement therapy (HRT), specifically estradiol, was associated with lower mortality in women aged 40 years or older (HR 0.77), with a similar but imprecise estimate for tibolone. These findings are consistent with prior reports showing benefits of estrogen maintenance on cardiovascular, bone, and metabolic health in the post-menopausal period^23^, but contrast with the trials of the Women’s Health Initiative (WHI), which did not support HRT for the prevention of cardiovascular and other chronic diseases^4,5^. Our estimates are, however, not directly comparable with the WHI. The trials tested oral conjugated equine estrogens, alone or combined with medroxyprogesterone acetate, in women aged 50–79 years who were on average more than a decade past menopause and mostly free of menopausal symptoms, with the aim of preventing chronic disease. Current practice, and most prescriptions captured here, instead involve 17β-estradiol, frequently administered transdermally and combined with micronized progesterone, and initiated near menopause for symptom relief. In WHI analyses stratified by age, women who started HRT at 50–59 years had lower all-cause mortality during the intervention phase^4^, consistent with the hypothesis that the timing of initiation matters.

A particularly notable finding was the association between vaccination with the varicella-zoster virus (VZV) vaccine and lower all-cause mortality. The lower mortality association observed for vaccines, and particularly the varicella-zoster virus (VZV) vaccine, aligns with recent evidence showing a decreased risk of developing neurodegenerative disease. Recent experiments have shown that both the live attenuated and recombinant shingles vaccines significantly reduce the subsequent risk of developing dementia, with the recombinant formulation yielding a substantial increase in dementia-free time^24–26^. However, recent reports suggest that these results were not reproduced in a much larger cohort^27,28^, leaving the scientific question open. In our data, the Shingles vaccine association (pooled HR 0.44) partly extended to deaths from external causes and appeared soon after vaccination, and was attenuated against Td/Tdap vaccine recipients in a target trial emulation (HR 0.85, 0.70– 1.03). It was, however, robust to adjustment for comorbidity, lifestyle and self-rated health, and was maintained against unvaccinated individuals and pneumococcal vaccine recipients, suggesting that healthy-user selection contributes to, but may not fully explain, it. Metformin users did not show lower mortality than non-users in our data, consistent with reports that metformin may not extend lifespan compared with non-diabetic controls^29,30^. Likewise, we do not see a lower mortality association of SGLT2 inhibitors, although pleiotropic beneficial effects on health have been reported^31,32^. Interestingly, statins and lipid modifying agents’ mortality association were largely neutral; yet RCTs have shown a benefit of statins to reduce all-cause mortality^19^. This illustrates that, when treatment is targeted at higher-risk individuals, this design can miss true benefits. Lastly, despite interest in the field^33^, our dataset was not well suited to study the association between supplements (such as Vitamin D, or Omega-3) and all-cause mortality, as these are usually prescribed OTC and poorly recorded in EHR, while also being potentially confounded by indication (osteoporosis, frailty).

Future research may pivot from evaluating purely survival-based endpoints to assessing broader metrics of healthspan^34^. While decreasing mortality is a critical medical objective, understanding how these medications influence the accumulation of chronic diseases and functional decline over time is equally paramount. Subsequent longitudinal studies may investigate the temporal trajectories of comorbidities, or intrinsic capacity^35^ and frailty in patients prescribed these agents. Further work may study the effect of these drugs on biological age^36^. Determining whether therapies like HRT, and the zoster vaccine are also associated with lower morbidity and higher functional capacity in old age will be essential to determine the potential benefit of these drugs.

There are several other limitations to this study. First, patients were considered continuously exposed following their first prescription, reflecting an intent-to-treat^37^ (ITT) analog approach. This assumption may inflate lower mortality associations for one-off preventive interventions such as vaccines and antimalarial prophylaxis, while separately biasing acute short-course drugs such as antibiotics. For NHANES since medication data was only available at baseline, the time-to-drug was set to 0. Thus, NHANES contributed a baseline-exposure (prevalent-user) analysis, which is more susceptible to immortal-time and prevalent-user biases. Throughout, we describe medications as associated with lower mortality; given the observational design, these associations should not be interpreted as causal effects. In addition, some drug exposure may have been missed including medications bought over the counter, or given outside the recording system (for example, vaccinations given in pharmacies). Furthermore, the integrated healthcare databases carry structural and clinical heterogeneity. The six cohorts encompass fundamentally distinct data capture environments, spanning routine primary care to tertiary hospital settings. For instance, datasets such as the UK Biobank and the JMDC predominantly capture community-based, general practice (GP) outpatient prescriptions. In contrast, comprehensive EHR systems like CDARS and FinnGen incorporate a substantial volume of inpatient, hospital-administered therapeutics. Because JMDC is substantially larger than the other cohorts and NHANES captured only baseline (prevalent) medication use, both were excluded from the pooled estimates and are reported at the cohort level only. Lastly, target trial emulation was performed in a single cohort (AOU) with limited follow-up. We also attempted an emulation for estradiol, among women aged 45–59 years initiating systemic estradiol compared with initiators of vaginal estrogen, but the number of deaths in this age range was too low for an informative estimate.

In conclusion, this multinational atlas maps the associations between prescription medications and all-cause mortality across six cohorts on three continents. Most associations reflect the clinical context in which drugs are prescribed, but a small number of preventive medications, most notably the VZV vaccine, showed consistent inverse associations that were robust to conventional adjustment and, for the VZV vaccine, largely maintained in a target trial emulation, although attenuation against a comparable vaccine group suggests that healthy-user selection contributes to them. These signals warrant confirmation in randomized trials. Together with the cohort-level and pooled estimates made available with this work, these results provide a resource for prioritizing candidates for drug repurposing in ageing research and for designing confirmatory studies.

## METHODS

### Study Design and Data Sources

We conducted a multinational, retrospective, longitudinal cohort study using RWD from six independent, large-scale population biobanks and EHR databases spanning three continents. The integrated data sources from the United States included the AOU and the NHANES. European cohorts comprised the UK Biobank and the FinnGen study. Asian cohorts included the CDARS, representing the territory-wide public healthcare system of Hong Kong, alongside the JMDC claims database from Japan. A federated analytical approach was employed wherein standardized analytical scripts were executed locally within each cohort’s secure environment, and summary-level statistics were subsequently meta-analyzed. The analysis plan was not preregistered. This work complies with all relevant ethical regulations. A full list of the ethics boards that approved the study protocols is provided at the end of this section.

The AOU^10^ is a highly diverse United States initiative that integrates electronic health records, survey responses, and biospecimen data. It specifically focuses on historically underrepresented populations, providing comprehensive longitudinal health tracking for over 500,000 participants. The medication screen used Curated Data Repository (CDR) release R2025Q4R6. The NHANES^11^ provides a nationally representative US sample through extensive clinical, nutritional, and laboratory examinations. These cross-sectional baseline assessments are subsequently linked to longitudinal Medicare and mortality records to evaluate long-term health outcomes. The cohort is ∼100,000 subjects, with ∼34,000 >= 40yo.

The UKBB^12^ is a large prospective European cohort comprising approximately 500,000 adults who were aged 40 to 69 years at the time of recruitment. Medication data was available for around 200,000 individuals. It features deep phenotypic profiling, including genetic, lifestyle, and physical measurements, comprehensively linked to ongoing national electronic health records. The FINNGEN^13^ study (release 13) is a nationwide initiative that combines comprehensive genomic profiles with decades of longitudinal electronic health registry data for approximately 500,000 Finnish citizens. 350,000 individuals with medication and mortality available were included in the study.

The CDARS^14^ is a territory-wide EHR database managed by the Hospital Authority (HA) of Hong Kong. The HA is the sole public healthcare provider that manages 43 hospitals and institutions and 123 outpatient clinics in Hong Kong, covering >80% of all hospital admissions. The CDARS contains data on demographics, disease diagnoses, drug prescriptions, admissions, and laboratory test results. For the present analysis, we included a random cohort of approximately 628,000 older adults aged ≥60 years who had at least one admission record at in-patient, out-patient, or accident and emergency services between January 1, 2010, and December 31, 2018, with follow-up until December 31, 2025. The JMDC^15^ is a nationwide, anonymized claims database from health insurance associations for large- and medium-sized company employees and their dependent family members aged <75 years. The data include demographics, inpatient and outpatient medical claims, pharmacy dispensing records, and annual health checkup data, covering >20 million insured individuals with a study period from January 1, 2005, to February 1, 2025.

### Study Population

The base population for each cohort included adult individuals aged 40yo or older. The index date (“time-zero”) was defined as the date of study entry or baseline assessment within each respective cohort, while in FinnGen the index date was defined as the date of first prescription record. Individuals with missing data for critical baseline demographic variables, such as age and sex, or whose death was recorded on or before their index date were excluded from the analytical cohort.

### Drug curation and standardization

Drug exposure data were curated and harmonised through a combination of automated coding and manual review, tailored to the source nomenclature of each cohort. For UKBB, AOU, NHANES, and CDARS, drug names were extracted from free-text or proprietary fields and manually curated to resolve brand-name/generic equivalences, spelling variants, and multi-ingredient product names, mapping each entry to a single active principle. For JMDC, standardised drug names were already available in the source data and required no additional curation. For FINNGEN, drugs were identified and classified using Anatomical Therapeutic Chemical (ATC) codes. Following cohort-level curation, all drug names were harmonised across the six cohorts into a single unified nomenclature based on active ingredient, enabling direct cross-cohort comparisons of individual drugs and drug classes. To ensure analytical stability, only drugs prescribed to more than 100 subjects within a given cohort were retained for analysis.

### Exposure Definition and Time-Dependent Ascertainment

The primary exposure of interest was the utilization of prescription medications. To mitigate immortal time bias, prescription drug exposure was modeled as a time-dependent covariate. Follow-up time for all individuals contributed to the unexposed risk period until the date of their first incident prescription for a given drug or drug class. From the date of the first prescription onward, their subsequent follow-up time was transitioned to the exposed risk period. Individuals whose first recorded prescription preceded the index date were classified as exposed from the index date onward (prevalent users). Patients were considered continuously exposed following their initial prescription, reflecting an intent-to-treat analog approach. Dose and duration of use were not modelled, and each drug was analysed separately against all individuals not (yet) exposed to it, without adjustment for co-medication. For NHANES, since medication data was only available at baseline, the time-to-drug was set to 0, and the same model was used otherwise. JMDC patients were censored after age 75.

### Outcome Ascertainment

The primary endpoint was all-cause mortality. Mortality status and the exact date of death were ascertained through linkage to national and regional death registries specific to each cohort. These included the National Death Index for the United States cohorts, the NHS Information Centre and central registries for the European cohorts, and the Hong Kong

Death Registry for the CDARS. For JMDC, death information was recorded in enrollment data. Follow-up continued until the date of death, loss to follow-up or administrative censoring (end of study period), whichever occurred first. In FinnGen, the underlying cause of death was obtained from the national cause-of-death registry (Statistics Finland), coded according to ICD-10.

### Covariates

To control for confounding by indication and baseline health status, a comprehensive set of covariates was extracted and harmonized across the six databases. Demographic variables included age at the index date, biological sex and enrollment date. Comorbidity burden was quantified using the Charlson Comorbidity Index (CCI), which was calculated for each patient using baseline diagnostic codes recorded before or at the index date. In UKBB, for which individual-level data could not be re-accessed at the time of this analysis, the CCI was approximated by the sum of baseline circulatory disease, diabetes and cancer (range 0–3). Furthermore, where available within specific cohorts, lifestyle and socioeconomic determinants were extracted. These included BMI, smoking status (categorized as never, former, or current), and harmonized indices of socioeconomic status such as the Townsend index (UKBB only). When the data was missing for whole variables (for ex. BMI, and smoking status in CDARS) the respective cohorts were excluded from the sub-analyses using these variables. In general the Cox was fitted only on complete cases. In AOU, self-rated general health (excellent, very good, good, fair or poor) from the participant surveys was additionally available and was used in a sensitivity analysis adjusting Model 1 for this variable.

### Time-Dependent Survival Analysis

To evaluate the association between prescription medication and all-cause mortality while rigorously accounting for immortal time bias, we employed a time-dependent covariate survival analysis framework utilizing Cox proportional hazards models (R survival package, tmerge, coxph functions). The primary timescale for the survival analysis was defined as the time on study, calculated from the date of cohort enrollment to the date of death or right-censoring. Drug exposure was inherently modeled as a dynamic variable to reflect the realities of longitudinal patient care. For each evaluated medication, the specific time to first exposure was calculated in years as the interval between the baseline enrollment date and the first recorded date of prescription issuance.

To execute this, we utilized a counting process approach, restructuring patient observation periods into distinct start and stop intervals. Medication use was incorporated into the Cox proportional hazards models as a binary, time-varying covariate. Under this analytical framework, an individual contributed unexposed person-time to the baseline cohort risk pool until the exact date of their first prescription, after which their subsequent follow-up time was classified as exposed. To ensure the statistical stability and reliability of our hazard ratio estimates, a Cox model was only computed for a specific medication if there were at least 15 observed mortality events among the actively exposed patient subset. For incident cancer, diabetes and heart disease in AOU (Figure 4A), the same time-dependent Cox model, adjusted for age and sex, was fitted with the first recorded diagnosis as the event, excluding individuals diagnosed before enrollment; adjusted cumulative event curves were derived for a woman of mean age with and without exposure.

### Hierarchical Confounder Adjustment and Stratification

We applied a progressively stringent series of adjustment models: “Model 1” was adjusted for age at enrollment, sex, and the year of enrollment, “Model 2” was further adjusted for CCI, and “Model 3” further incorporated smoking status, body mass index, and socioeconomic status. We also conducted stratified analyses to evaluate medication-mortality associations within distinct, clinically relevant subpopulations. Independent time-dependent Cox models performed across patient strata defined by baseline circulatory disease (I00–I99), cancer (C00–C99), diabetes (E08-E13), and smoking status, as well as within an explicitly healthy sub-cohort entirely free of these major underlying comorbidities. Finally, to account for multiple testing, for each unique analytical scenario and model type, we applied Bonferroni correction, scaling the significance thresholds by the total number of evaluated drugs within that specific data subset.

### Meta-Analysis

To combine the cohort-specific hazard ratios, we conducted a random-effects meta-analysis across AOU, UKBB, FinnGen and CDARS. NHANES, in which medication use was only recorded at baseline (prevalent users), and JMDC, which comprised over 80% of all participants and would dominate pooled estimates, were not pooled but are reported at the cohort level. The meta-analysis was limited to medications that were evaluated in at least two of the pooled cohorts. We pooled these individual cohort estimates utilizing the generic inverse variance method implemented via the rma function in the R metafor package. Recognizing the inherent structural, demographic, and clinical heterogeneity across the global biobanks, we applied a random-effects model, estimating the between-study variance using the Restricted Maximum Likelihood (REML) method. Meta-analysis p-values were Bonferroni-corrected for the number of drugs pooled within each analysis (e.g., 317 drugs in the main Model 1 analysis).

### Active comparator analysis

To reduce confounding by indication and healthy-user bias for the medications with the strongest inverse associations in the primary screen, we compared each with one or more active comparators. Comparators were selected using the following criteria:

(i) shared indication or use context: the comparator is prescribed for the same condition or in the same preventive setting (e.g., other vaccines routinely recommended for adults of the same age; other systemic estrogen formulations used for menopausal symptoms);
(ii) comparable patient and prescriber profile: initiating either drug implies a similar level of healthcare engagement, so that healthy-user effects are expected to be shared;
(iii) sufficient data: ≥100 new users and ≥15 deaths in at least two cohorts.

Applying these criteria, the comparators were: influenza and pneumococcal vaccines for the VZV and DTP vaccines; estriol, tibolone and conjugated estrogens for estradiol. In a new-user active-comparator design, patients initiating drug A or drug B were followed from first initiation until death or end of follow-up, with Cox models estimating the HR for A versus B adjusted for age at initiation, sex, and time from cohort entry to initiation.

### Negative-control exposure and cause-specific mortality

To probe healthy-user bias, we used typhoid vaccine as a negative-control exposure for atovaquone/proguanil: it is prescribed to the same population of international travelers but has no plausible effect on all-cause mortality in the populations studied. Its association with mortality was estimated with the same time-dependent Cox models as all other drugs. The DTP vaccine was interpreted in the same way for other adult vaccines. In FinnGen, we additionally estimated cause-specific mortality for the Shingles vaccine and atovaquone/proguanil according to the underlying cause of death: external causes (ICD-10 V01–Y98), including falls (W00–W19), transport accidents (V01–V99) and suicide (X60–X84); cancer (C00–D48); and circulatory disease (I00–I99). For each cause, deaths from that cause were treated as events and deaths from other causes were censored at the date of death (cause-specific hazards), using Model 1. External causes served as negative-control outcomes that neither drug can plausibly prevent. Estimates based on fewer than five deaths among exposed individuals are not reported.

### Target trial emulation of VZV vaccination

To test the VZV vaccine association with an explicit time zero, we emulated a target trial^18^ in AOU, using Curated Data Repository (CDR) release R2025Q4R6. Time zero could fall between January 1, 2017 and the data cutoff (December 31, 2024). Eligible individuals were aged 50–89 years, were enrolled in AOU, had at least one year of EHR history and at least one healthcare visit in the preceding two years, and had no prior record of zoster vaccination. Time zero was the date of the first recorded zoster vaccine dose (live attenuated or recombinant), identified from drug and procedure records. Only vaccinations after enrollment were included, to avoid immortal time between vaccination and enrollment. Follow-up continued until death or the end of data availability.

Three comparator groups were used. First, each vaccinee was matched on sex and birth year to up to five individuals who met the same eligibility criteria and were alive and unvaccinated on the vaccinee’s vaccination date (risk-set sampling, equivalent to a sequence of trials, one per vaccination day); matched comparators shared the vaccinee’s time zero. Second and third, in new-user active-comparator designs, vaccinees were compared with individuals receiving a pneumococcal vaccine or a tetanus-diphtheria vaccine (Td or Tdap) who had no zoster vaccination on or before that date, with time zero at the comparator vaccination and the same eligibility criteria. Individuals could contribute to both arms at different time zeros.

Confounding was addressed with propensity-score weights targeting the average treatment effect in the treated, truncated at the 99th percentile. Propensity scores included age, calendar year, the Charlson Comorbidity Index and major comorbidities, the number of healthcare visit days and of distinct drugs in the previous two years, prior influenza vaccination, prior travel vaccines or antimalarials, smoking, education, BMI and self-rated health, all measured before time zero; active-comparator models additionally included sex and prior pneumococcal and Td/Tdap vaccination. Balance was assessed with standardized mean differences (target <0.1). Weighted Cox models with robust standard errors clustered on individuals estimated intention-to-treat HRs for all-cause mortality. To detect selection at the time of vaccination, HRs were also estimated for 0–90 days, 91–365 days and more than one year after time zero, since a vaccine is unlikely to reduce mortality within weeks of administration^21^. Three-year cumulative mortality was estimated with weighted Kaplan– Meier estimators, with 95% CIs from 50 bootstrap resamples of matched sets or individuals. Sensitivity analyses omitted self-rated health from the propensity score, additionally adjusted the outcome model for covariates, and restricted the zoster arm to recombinant vaccine or to vaccinations not given on the same day as the comparator vaccine.

## Supporting information

Recode checklist

meta_analysis

results_by_cohort

## Data availability

Individual level data is only accessible through proper channel authorization in each cohort. We provide summary statistics for the meta-analysis, and results in individual cohorts. Cohort-level and pooled summary statistics are available in the results folder of the GitHub repository listed under Code availability.

## Code availability

The main function used to calculate the medication-mortality associations using a Time-Dependent Covariate (TDC) Cox model is provided with synthetic data on GitHub (https://github.com/perez-kevin/medication-mortality-biobank). Most results in this manuscript were derived using variations of this function, or as stated in the Methods section. Code for other analysis is also provided without synthetic data.

## Author contribution

K. P. conceived and designed the study. K. P., J. M. performed the statistical analyses, drafted the manuscript. K. T. contributed to data acquisition. A. G., J. J., C-L. C, A. A. provided access to and oversight of the cohort data. J. J., A. C., A.A. critically revised the manuscript. All authors approved the final version for submission.

## Acknowledgements

This research has been conducted using the UK Biobank Resource under application number 107065. We acknowledge All of Us participants for their contributions. We thank the National Institutes of Health’s All of Us Research Program for making available the participant data examined in this study. The All of Us Research Program is supported by the National Institutes of Health, Office of the Director: Regional Medical Centers: 1 OT2 OD026549; 1 OT2 OD026554; 1 OT2 OD026557; 1 OT2 OD026556; 1 OT2 OD026550; 1 OT2 OD 026552; 1 OT2 OD026553; 1 OT2 OD026548; 1 OT2 OD026551; 1 OT2 OD026555; IAA #: AOD 16037; Federally Qualified Health Centers: HHSN 263201600085U; Data and Research Center: 5 U2C OD023196; Biobank: 1 U24 OD023121; The Participant Center: U24 OD023176; Participant Technology Systems Center: 1 U24 OD023163; Communications and Engagement: 3 OT2 OD023205; 3 OT2 OD023206; and Community Partners: 1 OT2 OD025277; 3 OT2 OD025315; 1 OT2 OD025337; 1 OT2 OD025276. In addition, the All of Us Research Program would not be possible without the partnership of its participants. This study used publicly available data from the NHANES, conducted by the National Center for Health Statistics (NCHS), Centers for Disease Control and Prevention (CDC). This study used data from the JMDC Claims Database, provided by JMDC Inc. We thank the Hospital Authority of Hong Kong for providing access to the Clinical Data Analysis and Reporting System (CDARS) for research purposes. We want to acknowledge the participants and investigators of the FinnGen study (proposal index: F_2025_093). FinnGen is funded byBusiness Finland (grants HUS 4685/31/2016 and UH 4386/31/2016) and by industry partners (full list in Supplementary Note).

## Ethics statements

The study protocol was approved by the respective Institutional Review Boards and ethics committees overseeing each constituent cohort. The UK Biobank study was approved by the North West Multi-centre Research Ethics Committee (approval no. 11/NW/03820). All participants provided written informed consent for data collection, analysis and record linkage. For CDARS, the study protocol was approved by the Institutional Review Board of The University of Hong Kong/Hospital Authority Hong Kong West Cluster (Ref.: UW 24-347). The use of JMDC data was approved by JMDC Inc. As the CDARS and JMDC data were anonymized and no patient contact was involved, informed consent was not required in accordance with relevant local regulations. Study subjects in FinnGen provided informed consent for biobank research based on the Finnish Biobank Act, or were enrolled in study-specific cohorts collected prior to the Biobank Act and later transferred to Finnish biobanks under Fimea approval. The FinnGen study was approved by the Coordinating Ethics Committee of the Hospital District of Helsinki and Uusimaa (statement number HUS/990/2017), with additional study permits from the Finnish Institute for Health and Welfare, the Digital and Population Data Service Agency, the Social Insurance Institution, Findata, Statistics Finland, and the Finnish Registry for Kidney Diseases. Biobank access decisions for the samples and data used are listed in Supplementary Note. All of Us data were accessed through the Researcher Workbench under the Registered Tier, in accordance with the All of Us Data Use and Registration Agreement; per All of Us policy, analyses of de-identified Registered Tier data do not require separate institutional review board approval. NHANES data are publicly available and de-identified; the NHANES protocols were approved by the NCHS Research Ethics Review Board, and all participants provided written informed consent.

---

**Figure S1.**
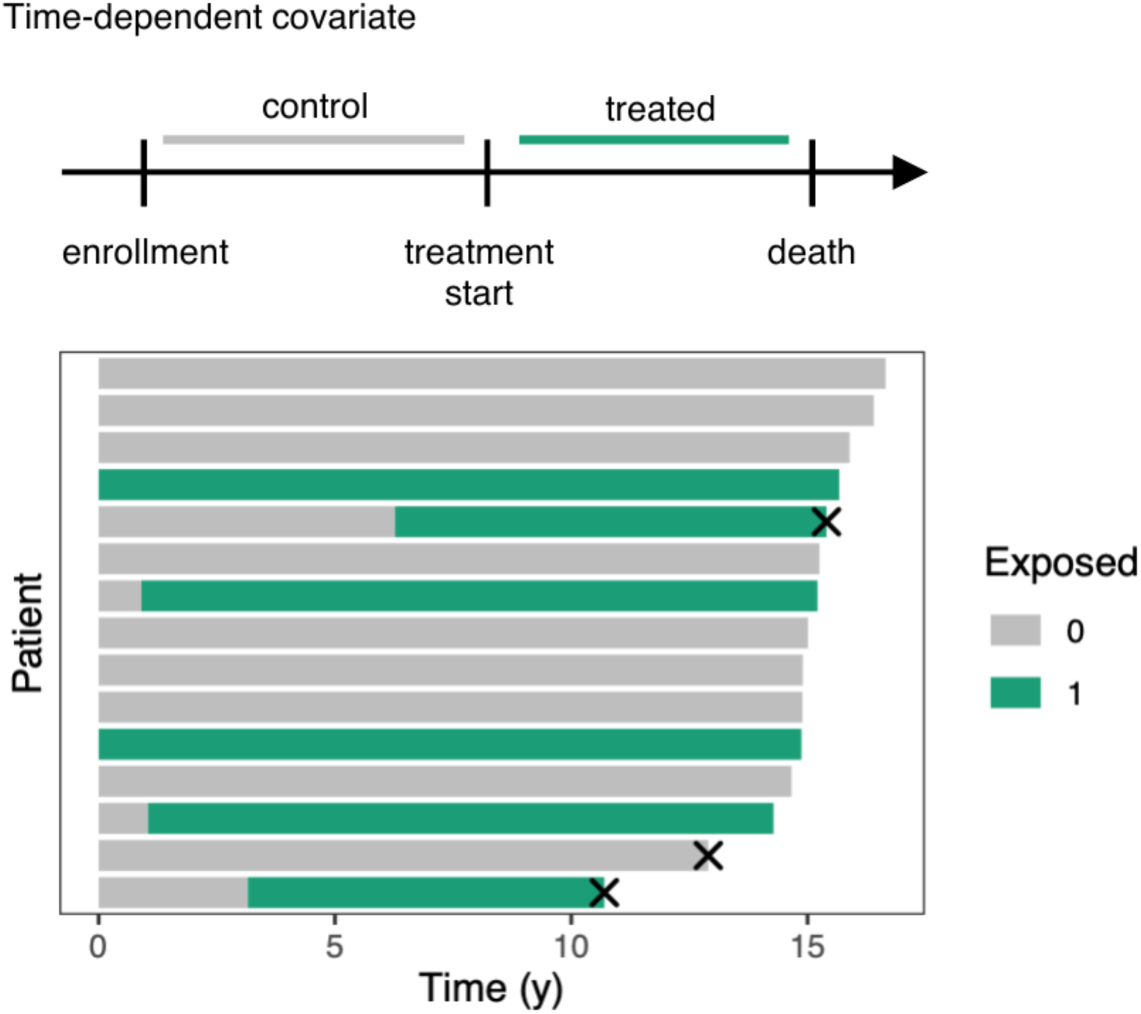
Illustration of the time-dependent covariate (TDC) analysis. Individuals contribute unexposed (control) person-time from enrollment until their first prescription, and exposed person-time from then until death or end of follow-up.

**Figure S2.**
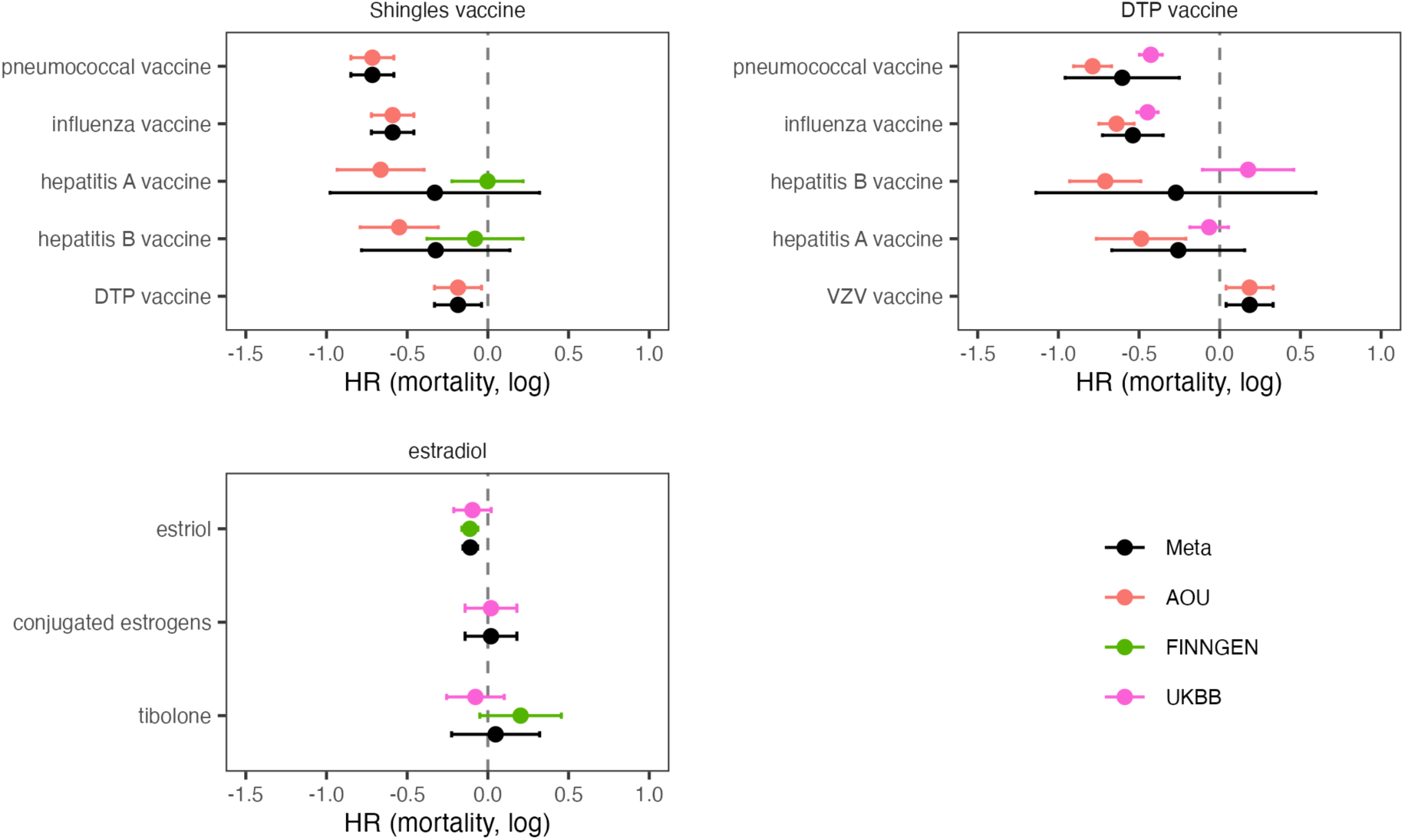
Active comparator analysis. Mortality log(HR) for the target drug (panel title) versus each comparator drug (y-axis), in a new-user active-comparator design adjusted for age at initiation, sex and time from cohort entry to initiation. Pooled (black) and cohort-specific estimates are shown.

**Figure S3.**
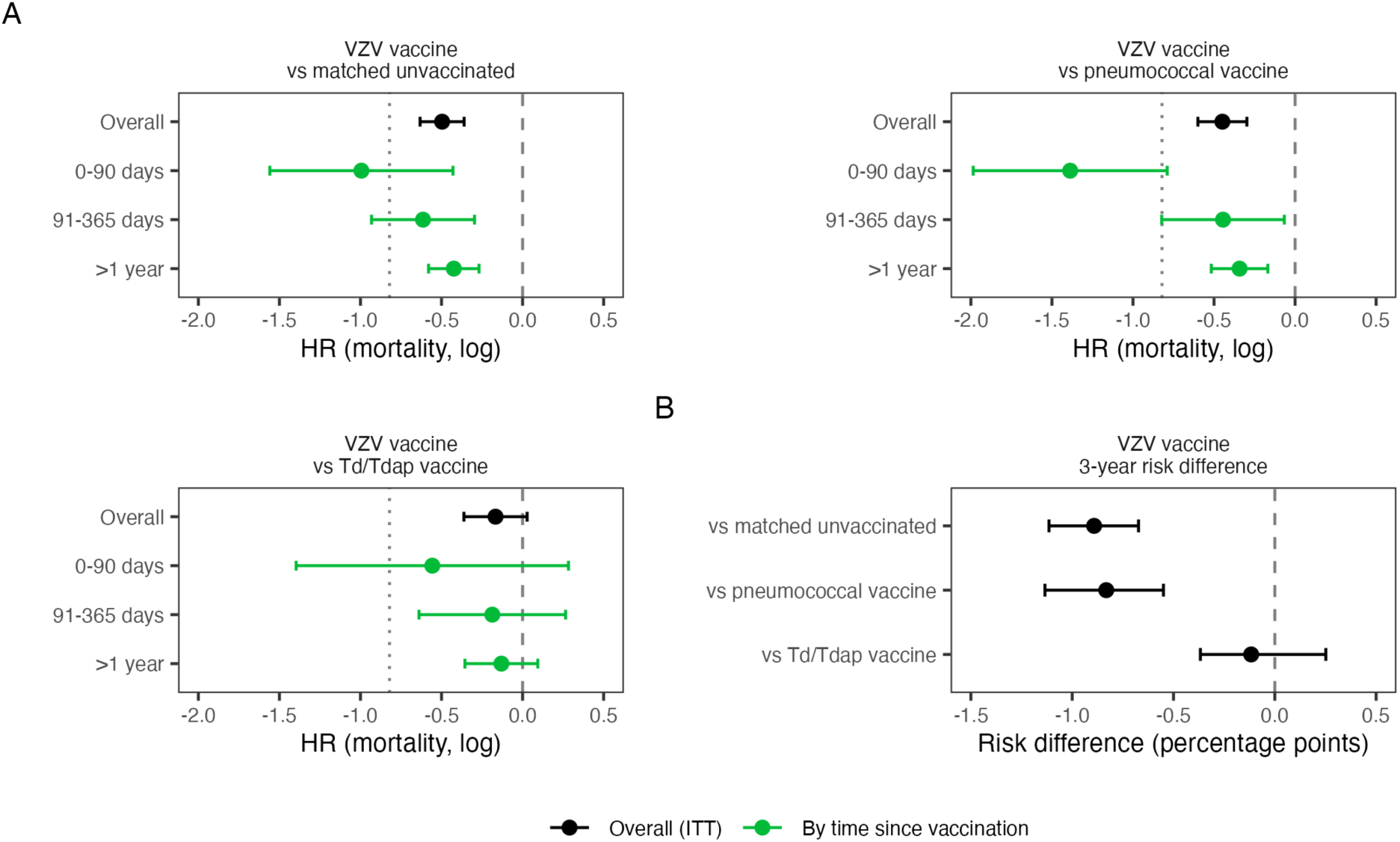
Target trial emulation of VZV vaccination in All of Us. (A) Mortality log(HR) (95% CI) among zoster vaccine recipients compared with matched unvaccinated individuals, pneumococcal vaccine recipients and Td/Tdap vaccine recipients, overall (intention-to-treat, propensity-score weighted including self-rated health; black) and by time since vaccination (green). The dotted line marks the pooled atlas estimate (Model 1, HR 0.44) and the dashed line no association. (B) Difference in 3-year cumulative mortality (percentage points, 95% bootstrap CI) from weighted Kaplan–Meier estimates.

**Figure S4.**
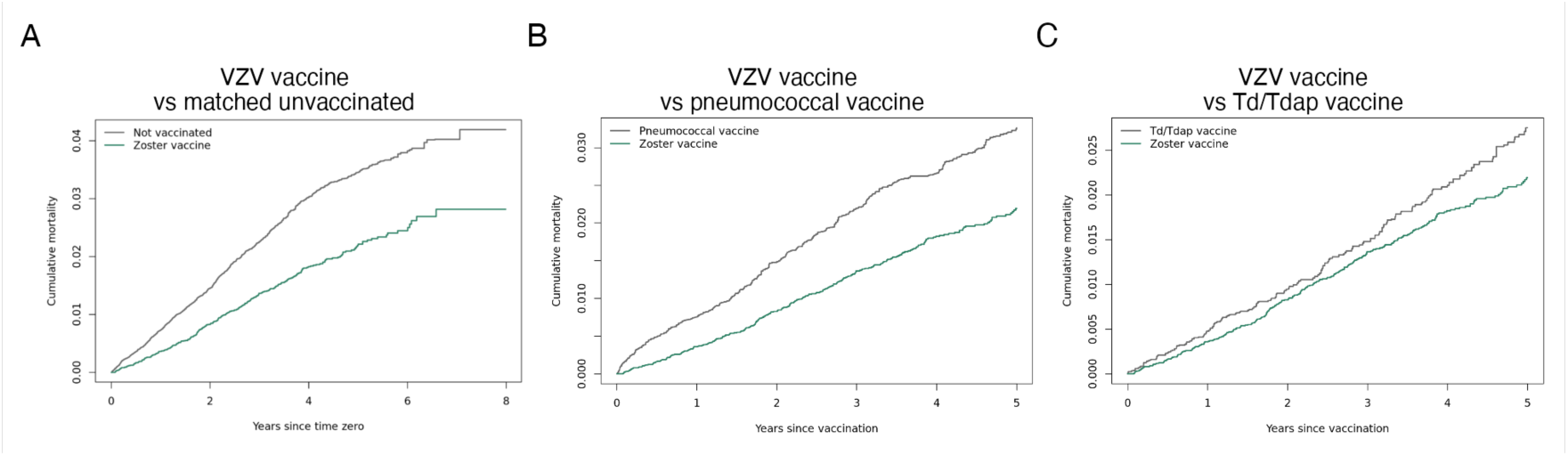
Weighted cumulative mortality after zoster vaccination in All of Us, compared with (A) matched unvaccinated individuals, (B) pneumococcal vaccine recipients and (C) Td/Tdap vaccine recipients. Time zero is the date of vaccination (in A, the vaccinee’s vaccination date for both groups).

