## Supplementary note for "A multinational pharmacoepidemiology atlas of prescription medication associations with all-cause mortality across six population cohorts"

### Acknowledgements

We want to acknowledge the participants and investigators of the FinnGen study. The FinnGen project is funded by two grants from Business Finland (HUS 4685/31/2016 and UH 4386/31/2016) and the following industry partners: AbbVie Inc., Alnylam Pharmaceuticals, Inc., AstraZeneca UK Ltd, Bayer AG, Biogen MA Inc., Boehringer Ingelheim International GmbH, Bristol Myers Squibb Inc. (and Celgene Corporation & Celgene International II Sàrl), Genentech Inc., GlaxoSmithKline Intellectual Property Development Ltd., Johnson&Johnson Innovative Medicine Inc., Maze Therapeutics Inc., Merck Sharp & Dohme LCC, Novartis AG, Pfizer Inc. and Sanofi US Services Inc. Following biobanks are acknowledged for delivering biobank samples to FinnGen: Auria Biobank (www.auria.fi/biopankki), THL Biobank (www.thl.fi/biobank), Helsinki Biobank (www.helsinginbiopankki.fi), Biobank Borealis of Northern Finland (https://www.ppshp.fi/Tutkimus-ja-opetus/Biopankki/Pages/Biobank-Borealis-briefly-in-English.aspx), Finnish Clinical Biobank Tampere (www.tays.fi/en-US/Research_and_development/Finnish_Clinical_Biobank_Tampere), Biobank of Eastern Finland (www.ita-suomenbiopankki.fi/en), Central Finland Biobank (www.ksshp.fi/fi-FI/Potilaalle/Biopankki), Finnish Red Cross Blood Service Biobank (www.veripalvelu.fi/verenluovutus/biopankkitoiminta), Terveystalo Biobank (www.terveystalo.com/fi/Yritystietoa/Terveystalo-Biopankki/Biopankki/) and Arctic Biobank (https://www.oulu.fi/en/university/faculties-and-units/faculty-medicine/northern-finland-birth-cohorts-and-arctic-biobank). All Finnish Biobanks are members of BBMRI.fi infrastructure (https://www.bbmri-eric.eu/national-nodes/finland/). Finnish Biobank Cooperative -FINBB (https://finbb.fi/) is the coordinator of BBMRI-ERIC operations in Finland. The Finnish biobank data can be accessed through the Fingenious® services (https://site.fingenious.fi/en/) managed by FINBB.

### Ethics statement

Study subjects in FinnGen provided informed consent for biobank research, based on the Finnish Biobank Act. Alternatively, separate research cohorts, collected prior the Finnish Biobank Act came into effect (in September 2013) and start of FinnGen (August 2017), were collected based on study-specific consents and later transferred to the Finnish biobanks after approval by Fimea (Finnish Medicines Agency), the National Supervisory Authority for Welfare and Health. Recruitment protocols followed the biobank protocols approved by Fimea. The Coordinating Ethics Committee of the Hospital District of Helsinki and Uusimaa (HUS) statement number for the FinnGen study is Nr HUS/990/2017.

The FinnGen study is approved by Finnish Institute for Health and Welfare (permit numbers: THL/2031/6.02.00/2017, THL/1101/5.05.00/2017, THL/341/6.02.00/2018, THL/2222/6.02.00/2018, THL/283/6.02.00/2019, THL/1721/5.05.00/2019 and THL/1524/5.05.00/2020), Digital and population data service agency (permit numbers: VRK43431/2017-3, VRK/6909/2018-3, VRK/4415/2019-3), the Social Insurance Institution (permit numbers: KELA 58/522/2017, KELA 131/522/2018, KELA 70/522/2019, KELA 98/522/2019, KELA 134/522/2019, KELA 138/522/2019, KELA 2/522/2020, KELA 16/522/2020), Findata permit numbers THL/2364/14.02/2020,

THL/4055/14.06.00/2020, THL/3433/14.06.00/2020, THL/4432/14.06/2020, THL/5189/14.06/2020, THL/5894/14.06.00/2020, THL/6619/14.06.00/2020, THL/209/14.06.00/2021, THL/688/14.06.00/2021, THL/1284/14.06.00/2021, THL/1965/14.06.00/2021, THL/5546/14.02.00/2020, THL/2658/14.06.00/2021, THL/4235/14.06.00/2021, THL/4990/14.02.00/2023 Statistics Finland (permit numbers: TK-53-1041-17 and TK/143/07.03.00/2020 (earlier TK-53-90-20) TK/1735/07.03.00/2021, TK/3112/07.03.00/2021) and Finnish Registry for Kidney Diseases permission/extract from the meeting minutes on 4th July 2019.

The Biobank Access Decisions for FinnGen samples and data utilized in FinnGen Data Freeze 13 include: THL Biobank BB2017_55, BB2017_111, BB2018_19, BB_2018_34, BB_2018_67, BB2018_71, BB2019_7, BB2019_8, BB2019_26, BB2020_1, BB2021_65, BB22-0025-A01, BB22-0025-A03, BB23-0222-A01, BB22-0025-A04, BB22-0025-A06, BB22-0025-A08, THLBB2024_30. Finnish Red Cross Blood Service Biobank 7.12.2017, 13.11.2023, 001-2023, Helsinki Biobank HUS/359/2017, HUS/248/2020, HUS/430/2021 §28, §29, HUS/150/2022 §12, §13, §14, §15, §16, §17, §18, §23, §58, §59, HUS/128/2023 §18, BB22-0025-A01, BB22-0025-A02, BB22-0025-A05, BB22-0025-A07, BB22-0025-A09, BB22-0025-A10, BB22-0025-A03, BB23-0222-A01, BB22-0025-A04, BB22-0025-A06, BB22-0025-A08, Amendment_BB22-0025-A05, Decision allowing to continue data processing until 31st Aug 2027: BB_2021-0140, HUS/150/2022 §12, BB_2021-0139, HUS/150/2022 §13, BB_2021-0161,HUS/150/2022 §14, BB_2021-0164, HUS/150/2022 §15, BB_2021-0169, HUS/150/2022 §16, BB_2021-0170, HUS/150/2022 §17, BB_2021-0179, HUS/150/2022 §18, BB_2022-0262, HUS/150/2022 §58, BB22-0067, HUS/150/2022 §59, Auria Biobank AB17-5154 and amendment #1 (August 17 2020) and amendments BB_2021-0140, BB_2021-0156 (August 26 2021, Feb 2 2022), BB_2021-0169, BB_2021-0179, BB_2021-0161, AB20-5926 and amendment #1 (April 23 2020) and it´s modifications (Sep 22 2021), BB_2022-0262, BB_2022-0256, BB22-0025-A01, BB22-0025-A02, BB22-0025-A03, BB23-0222_A01, BB22-0025-A02, BB22-0025-A05, BB22-0025-A07, BB22-0025-A09, BB22-0025-A10, BB22-0025-A03, BB23-0222-A01, BB22-0025-A04, BB22-0025-A06, BB22-0025-A08, Decision allowing to continue data processing until 31st Aug 2027: AB20-5926, BB_2021-0140, BB_2021-0156, BB_2021-0161, BB_2021- 0161, BB_2021-0164, BB_2021-0169, BB_2021-0179, BB_2022-0262, Biobank Borealis of Northern Finland_2017_1013, 2021_5010, 2021_5010 Amendment, 2021_5018, 2021_5018 Amendment, 2021_5015, 2021_5015 Amendment, 2021_5015 Amendment_2, 2021_5023, 2021_5023 Amendment, 2021_5023 Amendment_2, 2021_5017, 2021_5017 Amendment, 2022_6001, 2022_6001 Amendment, 2022_6006 Amendment, 2022_6006 Amendment_2, BB22-0067, 2022_0262, 2022_0262 Amendment, BB22-0025-A01, BB22-0025-A02, BB22-0025-A05, BB22-0025-A07, BB22-0025-A09, BB22-0025-A10, BB22-0025-A03, BB23-0222-A01, BB22-0025-A04, BB22-0025-A06, BB22-0025-A08, Decision allowing to continue data processing until 31st Aug 2027: BB/2021/5015, BB/2021/5017, BB/2021/5018, BB/2021/5023, BB/2022/6006, BB/2022/6001, BB/2022-0262, BB/2021/5010, Biobank of Eastern Finland 1186/2018 and amendment 22§/2020, 53§/2021, 13§/2022, 14§/2022, 15§/2022, 27§/2022, 28§/2022, 29§/2022, 33§/2022, 35§/2022, 36§/2022, 37§/2022, 39§/2022, 7§/2023, 32§/2023, 33§/2023, 34§/2023, 35§/2023, 36§/2023, 37§/2023, 38§/2023, 39§/2023, 40§/2023, 41§/2023, BB22-0025-A01, BB22-0025-A02, BB22-0025-A05, BB22-0025-A07, BB22-0025-A09, BB22-0025-A10, BB22-0025-A03, BB23-0222-A01, BB22-0025-A04, BB22-0025-A06, BB22-0025-A08, Decision allowing to continue data processing until 31st Aug 2027: MO-BB_2021-0179-A0, MO-BB_2021-0156_PRE-A01, BB_2021-0140, MO-BB_2021-0170_PRE-A0, MO-BB_2021-0169-A01, MO-BB_2022-0256-A01, MO-BB_2021-0161-A01, MO-BB_2021-0161-A02, BB22-0067-A01, MO-BB_2022-0262-A0, Finnish Clinical Biobank Tampere MH0004 and amendments (21.02.2020 & 06.10.2020), BB2021-0140 8§/2021, 9§/2021, §9/2022, §10/2022, §12/2022, 13§/2022, §20/2022, §21/2022, §22/2022, §23/2022, 28§/2022, 29§/2022, 30§/2022, 31§/2022, 32§/2022, 38§/2022, 40§/2022, 42§/2022, 1§/2023, BB2021-0140, BB22-0025-A01, BB_2021-0161, BB22-0025-A02, BB22-0025-A05, BB22-0025-A07, BB22-0025-A09, BB22-0025-A10, BB22-0025-A03, BB23-0222-A01, BB22-0025-A04, BB22-0025-A06, BB22-0025-A08, Decision allowing to continue data processing until 31st Aug 2027: BB_2021- 0140, BB_ 2021- 0161, BB_ 2021- 0179, BB_ 2021- 0156, BB_ 2021- 0169, BB_ 2021- 0170, BB22- 0067- A01, Central Finland Biobank 1-2017, BB_2021-0169, BB_2021-0179, BB_2022-0256, BB_2022-0262, Decision allowing to continue data processing until 31st Aug 2027 for projects: BB_2021-0179, BB22-0067,BB_2022-0262, BB_2021-0170, BB_2021-0164, BB_2021-0161, and BB_2021-0169, BB22-0025-A01, BB22-0025-A02, BB22-0025-A05, BB22-0025-A07, BB22-0025-A09, BB22-0025-A10, BB22-0025-A03, BB23-0222-A01, BB22-0025-A04, BB22-0025-A06, BB22-0025-A08, Terveystalo Biobank STB 2018001 and amendment 25th Aug 2020, Finnish Hematological Registry and Clinical Biobank decision 18th June 2021, Amendment 2nd January 2024 and Arctic biobank P0844: ARC_2021_1001, ARC_2023_3003 (BB22-0025-A01), BB22-0025-A03, BB23-0222-A01, BB22-0025-A04, BB22-0025-A06, BB22-0025-A08.
